# Have the COVID-19 Pandemic and Lockdown Affected Children’s Mental Health in the Long Term? A Repeated Cross-Sectional Study

**DOI:** 10.1101/2022.05.10.22272976

**Authors:** Manas Pustake, Sushant Mane, Mohammad Arfat Ganiyani, Sayan Mukherjee, Misba Sayed, Varada Mithbavkar, Zaid Memon, Abdus Samad Momin, Krishna Deshmukh, Ayush Chordia, Sabyasachi Parida, Ajit Bhagwat, Alan Johnson, Deepankar Varma, Sanket Warghade

## Abstract

**Objective:** The study aimed to evaluate the impact of the COVID-19 pandemic on levels of anxiety and depressive symptoms in children and adolescents.

**Design:** Cross-sectional surveys were carried out on the mental health of children; one survey was conducted before the COVID-19 pandemic and one into the pandemic, 15 months after the implementation of lockdown, social distancing, and school closures. Demographic data and COVID-19 pandemic-related data were collected from specific parent-report and self-report questionnaires.

**Participants:** Participants included children and adolescents between ages 6-16 years, attending a tertiary care hospital without any diagnosed major psychiatric disorder or chronic disorder.

**Analysis:** Data was collected at two points (before the COVID-19 pandemic and during it) and compared. Levels of anxiety and depressive symptoms were compared and tested for statistically significant differences between these two points using appropriate statistical tests. Regression models were constructed to predict the factors affecting increased anxiety levels and depressive symptoms in the COVID-19 period.

**Results:** 832 and 1255 children/adolescents were included in the study during the pre-COVID-19 and COVID-19 times, respectively. The median age of the participants was 10 years [Interquartile Range (IQR) = 4 years). The median (IQR) Spence Children’s Anxiety Scale score was 24 (12) at the pre-COVID-19 point and 31 (13) during the COVID-19 pandemic (p<0.001, r=-0.27). 11% and 16% of children reported being depressed at these two-time points, respectively (p=0.004, φ_c_=-0.063). Regression analysis showed that many factors, including the duration of smartphone use, female gender, and only child status, were associated with increased anxiety or depression levels.

**Conclusion:** A large proportion of children had elevated anxiety and depressive symptoms during the pandemic relative to before the pandemic, suggesting a need for measures to engage children in healthy habits to protect children’s mental health and continuous monitoring of children during such scenarios.

**STRENGTHS AND LIMITATIONS:**

- With the availability of pre-pandemic data, the repeated cross-sectional study design allowed us to compare the anxiety symptoms and prevalence of depression in children and adolescents during and before the COVID-19 lockdown and school closures.
- The study is one of the few studies from low-to-middle income countries on this topic with large sample size.
- The data was collected hospital setting, and all of the participants were attending a hospital, which could have resulted in a sampling bias. Although it is a tertiary care hospital, all of the patients included in the study came to us for primary care and were not referred.
- We used standardised scales that are usually used for screening and evaluation purposes and not for diagnostic purposes.
- We were unable to perform a longitudinal study with a follow-up that would offer clear evidence of any fluctuation in mental health during the course of the pandemic.

## INTRODUCTION

Mental health is viewed as the most crucial parameter for a high quality of life,[1]. Mentally healthy children can carry their enthusiasm and self-confidence into adulthood, giving them the ability to deal with hardships.

The COVID-19 pandemic resulted in the first-ever long-term closure of schools from March 2020 to October 2021,[2,3]. The COVID-19 pandemic and its subsequent containment measures, such as stay-at-home orders and quarantine, have had a detrimental effect on the mental health of individuals of all ages,[4]. In India, schools were closed from the beginning of the pandemic, with curricula transferred to an online platform within a month. During this period, a strict lockdown with stay-at-home orders was in place. Subsequently, from November 2020 to February 2021, recreational activities such as playgrounds and sports were opened in a phased manner. Then came another lockdown due to the second wave with strict stay-at-home orders, restricting all outdoor activities once again. The schools remained completely online during this entire time,[5].

The present pandemic represents a novel, complex, and multifaceted psychosocial stressor affecting children and adolescents’ mental health,[6]. In the wake of the closure of schools, social interactions and recreational activities outside of the home were highly restricted. Playgrounds, parks, turfs, swimming pools, and dance classes were closed,[7]. A lack of these activities caused by containment measures may affect children’s psychological well-being, particularly of vulnerable children,[8]. Parental stress might also have contributed to these children’s behavioural and psychological problems,[9]. Moreover, physical inactivity, dietary imbalance, and poor sleeping habits probably have contributed to exacerbating this problem, notably among impoverished and marginalised children and adolescents,[10–12].

The majority of primary evidence of the impact of flu and SARS/MERS outbreaks on mental health comes from cross-sectional study designs. These studies report significant mental health-related consequences such as psychological distress, anxiety, depression, and fear,[13–15]. There are rapidly accumulating repeated cross-sectional and longitudinal studies, including studies on children,[14,16–25]. Though there is much literature on the psychological impacts of the pandemic, in a living systematic review of ample abstracts on this topic, only a handful of abstracts were able to offer robust data on it, with very few addressing the impact on children,[26].

The present study is one of the few studies from a low-to-middle-income country. Besides this, the fact that the study was conducted in the country’s worst-affected city adds to our understanding of the impact on children’s mental health. The present study design aimed to compare symptoms of anxiety and depression before and during the pandemic. To broaden our approach, as a secondary aim, we also tried to look at the factors significantly associated with anxiety levels and depressive symptoms during the pandemic, such as sociodemographic factors and behavioural factors.

## MATERIALS AND METHODS

### Study Design

This study was a repeated cross-sectional study in which we included children and adolescents at two different points in time. The first sample point was between February 13, 2020, and March 5, 2020, before India’s countrywide strict lockdown and school closure. The second sample point was between May 1, 2021, and July 7, 2021. The initial data collection was carried out in the context of another cross-sectional study designed in similar settings. Due to the pandemic and lockdown state, the recruitment of subjects was temporarily suspended, and a decision was made to study the impact of lockdown on the population already participating. As a result, the final data collection was done during the second wave of COVID-19 lockdown, when social restrictions and school closures were in place for 15 months. [Physical schools were strictly closed in India from March 2020 to September 2021, shifting their routine curriculum to an online platform.] The data was collected offline through in-person interviews with observing COVID-19 appropriate behaviour.

### Study Setting and Participants

The study was undertaken at a tertiary health centre in Mumbai, India. All children in the age group 6-16 years visiting the outpatient and in-patient department of the paediatrics department for general consultation, immunisation, or refilling for medications were considered eligible for the study. Children who were previously diagnosed with any psychiatric disorder or other major chronic disorder admitted for a serious illness or used the medication that may affect the study results, such as atypical antipsychotics, were excluded. Parents or children/adolescents (as applicable) were asked to fill out the questionnaires on behalf of their children with the help of one of the investigators. Parent-proxy data was collected for children under the age of 11, and older children were tested with self-reports. However, considering the education, literacy, and ability to understand and respond to questions, ad-hoc decisions were taken to change to a parent interview or read out the questions to the parent/child in an interview format, if necessary. The questions were read with the items in the first-person form if they were read aloud. The investigators were highly trained medical students (final year or interns) who actively participated in the meeting or translation process by discussing the scales and questionnaire. Responses only from parents who had been with their child for more than six months before data collection were collected.

### Ethical Declaration

The study was approved by the institutional ethics committee of Grant Government Medical College and Sir JJ Group of Hospitals, Mumbai (Document Nos. IEC/Pharm/RP/387/Mar/2021 and IEC/Pharm/ICMR/59/Feb/2020). Children were recruited for the study only after their guardian gave written informed consent, and the children themselves gave assent for participation in the study. Children who had higher than cut-off scores in the screening tests were referred to the psychiatry department for further evaluation. Anthropometric parameters (viz. height and weight) were not measured because doing so could have increased their risk of exposure to COVID-19, so appropriate social distancing was maintained.

### Measurements

The questionnaires used in this study were primarily divided into four components.

1. **Case Record Form:** It was a form containing concise basic information about the socio-economic and demographic variables of the children and their families (like age, education, type of family, address, occupation, income, etc.). It also consisted of general questions like smartphone use, the child’s academic performance (in grades), recreational activity such as outdoor sports, and activity/exercise. The form and additional details regarding the definitions of the terms used in this study can be found in the <u>supplementary materials. (Supplementary files 1 and 2).</u>
2. **The Spence Child Anxiety Scale (SCAS)**,[27]: Anxiety levels were measured using the SCAS. It is a scale that uses response options, scaled using a 4-point Likert scale. It is used widely for assessing the anxiety levels of children and adolescents. This scale has 44 items, 38 of which reflect anxiety symptoms and six of which are positive filler items to avoid negative response bias. Within the questionnaire, items are assigned at random. Children are asked to report the frequency with which they encounter each symptom on a four-point scale: never (0), sometimes (1), often (2), and always (3). The cut-off score for significant anxiety levels varies with age and gender. This scale has been validated and widely used in various research settings, including India, with an excellent internal consistency (Cronbach’s alpha= 0.93),[28–31]. One of the advantages of using this measure is the availability of a parent-reported version.
3. **Centre for Epidemiological Studies—Depression Scale for Children (CES-DC)**,[32]: This scale is a revised form of the Centre for Epidemiologic Studies Depression Scale. It is a depression inventory of 20 items with possible sum scores varying from 0 to 60. In children and adolescents, a score of ≥ 15 suggests the presence of depressive symptoms. The scale is found to have a high internal reliability (Cronbach’s alpha = 0.84) and test/retest reliability (r = 0.51). This scale has been validated and widely used in various research settings, including India,[28,33]. The availability of a parent-proxy version is one of the benefits of using this measure.
4. A **special COVID-19 pandemic and lockdown related questionnaire** was devised by our research team, which had the questions about the following– Whether any parent is involved in anti-epidemic work/essential jobs, degree of worry of the pandemic, implementation of precautionary measures by the child, whether a close relative/friend infected with COVID-19 recently, whether a close relative/friend died with COVID-19, whether the child is ever infected with COVID-19, whether the parents yell at/beat their child, etc. The questionnaire is available in the <u>supplemental file</u>. <u>(Supplementary file 3).</u> Our research team developed the questionnaire after conducting a thorough literature review (including preprints). Those parameters, which were significant in published studies, were included in the questionnaire. We drew on relevant findings from these studies. Some questions were also suggested by colleagues in our department who were conducting similar types of studies (related to the pandemic). The investigators read out the questions rather than allowing the participants to fill out the questionnaires for convenience and avoid heterogeneity in the collected data.

All these questionnaires were made available in three languages, viz. English, Hindi, and Marathi for patients to understand. The scales were either translated into native languages as described below or the translated and validated version in the native language was used, if available. The case record forms and COVID-19 related questionnaire were first designed in native languages and then translated to English. All the questionnaire components were used during both time points of the study except the COVID-19 related questionnaire, which was employed only during the COVID-19 lockdown.

### Translation

Three of the authors, who were fluent in all three languages, first translated the items from English to Marathi and Hindi (for SCAS and CES-DC). Another author produced a back-translation. The original was compared to the back-translated version, and minor differences were resolved. The translated scale was evaluated and approved by a panel of experts (two Psychiatrists and a Paediatrician).

### Data Analysis

After collecting data, the data was entered into a spreadsheet using Microsoft Office Excel 2019 (Microsoft Corporation, WA, USA). Identifiable data were not entered into the spreadsheet.

### Statistical Analysis

For categorical variables, data were reported as absolute numbers and percentages. Medians with interquartile range (IQR) were used to represent ordinal and numerical variables that did not follow a normal distribution (tested with the Kolmogorov-Smirnov test). The chi-square test (for qualitative variables), Mann-Whitney U test, and Kruskal– Wallis H test (for comparing mean ranks between two groups if the normality was rejected) were used to assess the differences between groups with continuous or ordinal variables. A p-value <0.05 (2-tailed) was considered statistically significant. As appropriate, effect sizes were described as Cramer’s V and r measures.

CES-DC scores were categorised into depressed or non-depressed using the cut-off value as CES-DC is a validated scale with a gender and age non-dependent cut-off of 15 points, indicating clinically significant depressive symptoms,[34]. The SCAS scores were analysed as scores for the following reasons – 1) SCAS has a cut-off that varies with age and gender, 2) non-availability of standardised T-score sheets for certain age groups in our study, and 3) As our primary objective was to study whether the pandemic has affected the anxiety levels in children, which was achievable with the use of scores only. Multiple linear regression and bivariate logistic regression analyses were used to assess the association between outcome variables (the reported level of anxiety and significant depressive symptoms, respectively) and potential predictors (use of a smartphone, friend or family infected with COVID-19, observing COVID-19 appropriate behaviour, etc.) while adjusting for other identified explanatory variables. The forward stepwise selection algorithm was used to run models, and variables in the model were screened based on significance levels of the Wald inclusion test statistic being less than 0.05. Correlation between numerical variables was performed using Pearson’s correlation test, and scatterplots were plotted. Median values and groupwise distribution of numerical data were depicted in the violin plots. Statistical analysis was performed using the IBM SPSS Statistics software (v26.0; IBM, Armonk, NY, USA).

### Patient and public involvement

Patients or the general public were not involved in our study’s design, conduct, reporting, or dissemination.

## RESULTS

### Sociodemographic characteristics

The study included 843 children enrolled during the pre-COVID-19 study period and 1285 children enrolled during the COVID-19 lockdown period. Due to incomplete questionnaire responses during the pre-COVID-19 and COVID-19 lockdown study periods, we excluded 11 and 30 enrolled children from the analysis, respectively. The participant flow diagram of the study is depicted in Figure 1. The sociodemographic characteristics of the participants are described in table 1 (Table 1). The median age of the children and adolescents who participated was 9 (3) years and 10 (4) years at the two points, respectively. Females constituted 53% of the total number of children and adolescents who participated. All the participants were Asian (Indians) by ethnicity. In our sample, children (age 6-11 years) were almost two-thirds of the total participants, whereas adolescents (age 12-16 years) constituted the remaining participants. Around 86% of the participants’ mothers were literate, while the fathers had a literacy rate of 94.5%. The majority of the participants resided in urban areas (84.6%) and were attending the outpatient department (77.8%) at the time of data collection. 87.26% of the data was reported by parent proxy during the pre-pandemic time, and 81.36% of data was parent-proxy during the pandemic time. We shifted from the child-report to the parent-report version for 286 (45.68%) adolescents. 43 (2.06%), 69 (3.3%), and 1975 (94.63%) children were attending the hospital for immunisation, refilling of medications and general consultation, respectively.

**Table 1:** Sociodemographic characteristics of the participants.

|  | **Pre-COVID-19**<br>**(N = 832)** | **During the lockdown (N = 1255)** | **p-value** |
| --- | --- | --- | --- |
| **Gender** |  |  | 0.39<sup>§</sup> |
| Male | 401 (48.20%) | 581 (46.29%) |  |
| Female | 431 (51.80%) | 674 (53.71%) |  |
| **Age [Median (IQR)]** | 9 (3) | 10 (4) | <0.001*† |
| Children (6-11 years) | 636 (76.4%) | 825 (65.7%) |  |
| Adolescents (12-16 years) | 196 (23.6%) | 430 (34.3%) |  |
| **Type of Family** |  |  | 0.452<sup>§</sup> |
| Nuclear | 606 (72.84%) | 940 (74.90%) |  |
| Extended | 209 (25.12%) | 296 (23.59%) |  |
| Single Parent | 17 (02.04%) | 19(01.51%) |  |
| **Only Child** |  |  | 0.710<sup>§</sup> |
| No | 750 (90.14%) | 1125 (89.64%) |  |
| Yes | 82 (09.86%) | 130 (10.36%) |  |
| **Education of Mother** |  |  | 0.879<sup>§</sup> |
| Illiterate | 118 (14.2%) | 188 (15%) |  |
| Primary School | 168 (20.2%) | 246 (19.6%) |  |
| Secondary School | 350 (42.1%) | 513 (40.9%) |  |
| High School And Above | 196 (23.6%) | 308 (24.5%) |  |
| **Education of Father** |  |  | 0.091<sup>§</sup> |
| Illiterate | 46 (5.5%) | 81 (6.5%) |  |
| Primary School | 189 (22.7%) | 286 (22.8%) |  |
| Secondary School | 282 (33.9%) | 364 (29.0%) |  |
| High School and Above | 315 (37.9 %) | 524 (41.8%) |  |
| **Report type** |  |  | **<0.001\*<sup>§</sup>** |
| Self-report | 106 (12.74%) | 234 (18.64%) |  |
| Parent-proxy | 726 (87.26%) | 1021 (81.36%) |  |
| **Academic Grades** |  |  | **<0.001\*<sup>§</sup>** |
| A Grade | 556 (66.83%) | 915 (72.91%) |  |
| B Grade | 226 (27.16%) | 235 (18.72%) |  |
| C or D Grade | 50 (06.01%) | 105 (08.37%) |  |
| **Family Income; Median (IQR)**<br>(Lakh INR per annum) | 5 (5) | 5 (4) | 0.284<sup>†</sup> |
| **Residence** |  |  | **0.028\*<sup>§</sup>** |
| Urban | 627 (75.36%) | 997 (79.44%) |  |
| Rural | 205 (24.64%) | 258 (20.56%) |  |
| **Engages in recreational activities?** |  |  | **<0.001\*<sup>§</sup>** |
| No | 266 (31.97%) | 539 (42.95%) |  |
| Yes | 566 (68.03%) | 716 (57.05%) |  |
| **Smartphone User** |  |  | **<0.001\*<sup>§</sup>** |
| No | 245 (29.45%) | 65 (05.18%) |  |
| Yes | 587 (70.55%) | 1190 (94.82%) |  |
| **Smartphone Usage; Median (IQR) (hours/day)** | 1 (1) | 4 (2) | **<0.001\*<sup>†</sup>** |
| **Inpatient/Outpatient** |  |  | 0.707<sup>§</sup> |
| Outpatient | 701 (84.25%) | 1065 (84.86%) |  |
| Inpatient | 131 (15.75%) | 190 (15.14%) |  |
| **\*Significant, <sup>§</sup>Chi square Test, <sup>†</sup>Mann-Whitney U Test** |  |  |  |

**Figure 1:**
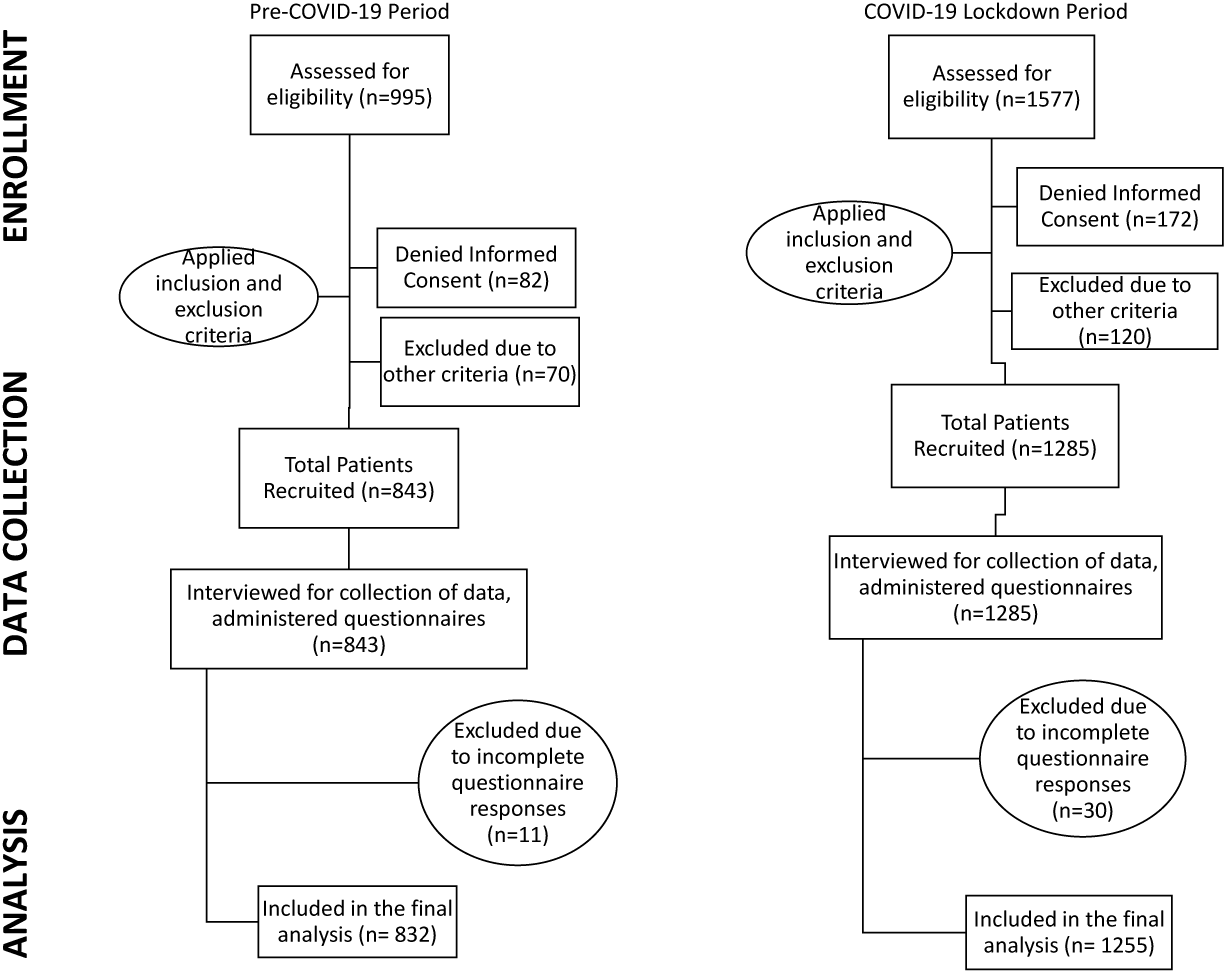
Participants flow diagram of patients visiting a tertiary care centre.

### Smartphone Usage

During the pre-COVID-19 period, the prevalence of smartphone use in children was 71%, which increased drastically to 95% during the pandemic (p<0.001). Moreover, the median (IQR) smartphone use per day increased from 1 (1) hour to 4 (2) hours during the pandemic (p=<0.001).

### Responses to COVID-19 related questionnaire

The responses to COVID-19 related questions are depicted in supplementary table 1. 25% of the children were worried “very much” about the pandemic, with 59% being “somewhat” worried and 16% being “not worried.” 80% of the participants reported observing COVID-19 appropriate behaviour, including the use of face masks and sanitisers. 5.4% of the children had a history of infection with the coronavirus in the past.

### Anxiety Levels of the participants before and during the pandemic

A comparison of anxiety levels using SCAS before and during the COVID-19 lockdown period is shown in Table 2 (Table 2). There was a statistically significant difference with small to medium effect in anxiety levels between these two groups (p<0.001, effect size r = -0.27). The median values and distributions of the SCAS scores are shown in violin plots with p-values from the Mann-Whitney U test. (Supplementary Figure 1; Panels A-C). SCAS Scores correlated weakly positively but statistically significantly with smartphone usage hours/day (Pearson’s r =0.209, p<0.001) and strongly but statistically significantly with CES-DC scores (r =0.680, p<0.001). Scatterplots of the same are depicted in figure 2 (panels A and C).

**Table 2:**
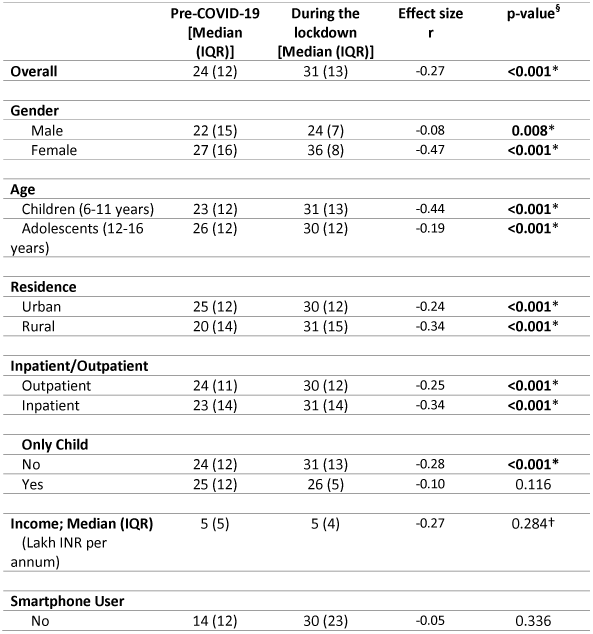
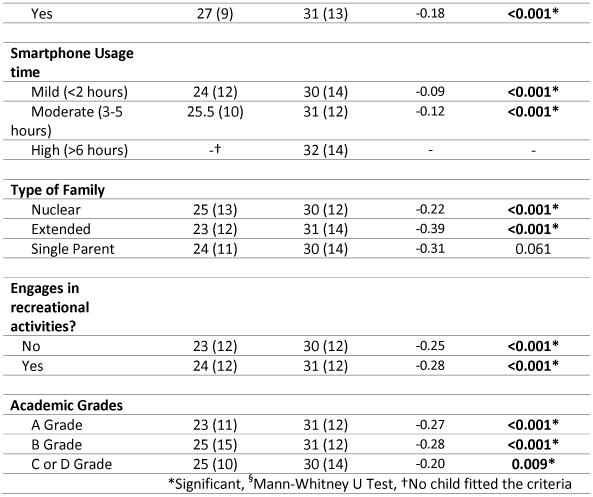
Anxiety using SCAS before and after the COVID-19 lockdown by the groups.

**Figure 2:**
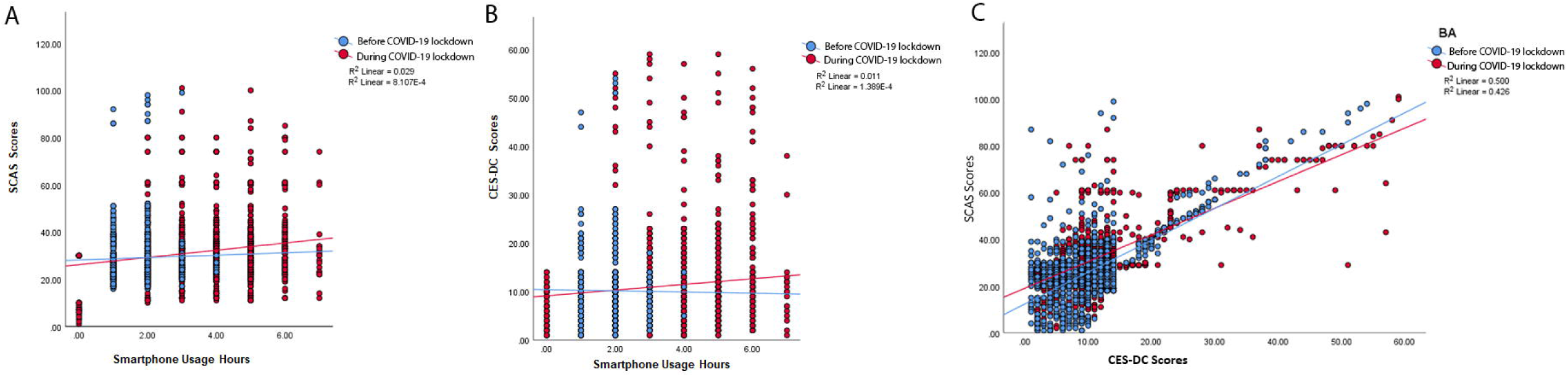
a) Scatterplot between SCAS scores and smartphone use per day (in hours) b) Scatterplot between CES-DC scores and smartphone use per day (in hours) c) Scatterplot between SCAS scores and CES-DC scores.

### Depressive symptoms in the participants before and during the pandemic

A comparison across depressive and non-depressive groups using CES-DC before and during the COVID-19 lockdown period is shown in Table 3 (Table 3). There was a statistically significant difference with a weak effect size between groups with depression levels above the cut-off score and those under the cut-off score before and during the COVID-19 lockdown period (p=0.004, φ_c_= 0.067). The distribution and median values of the CES-DC scores are shown in violin plots with p-values from the Mann-Whitney U test. (Supplementary Figure 1; Panels D-F) CES-DC Scores correlated very weakly positively but statistically significantly with smartphone usage hours/day (r =0.108, p=<0.001) (Figure 2; panels B and C).

**Table 3:**
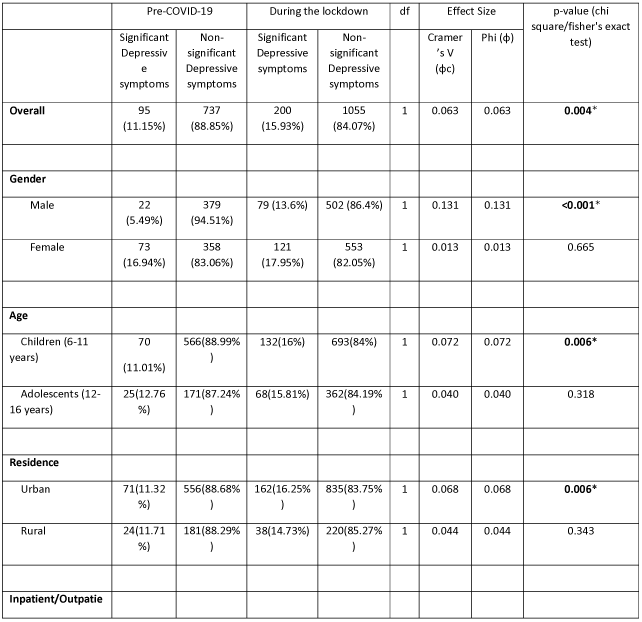
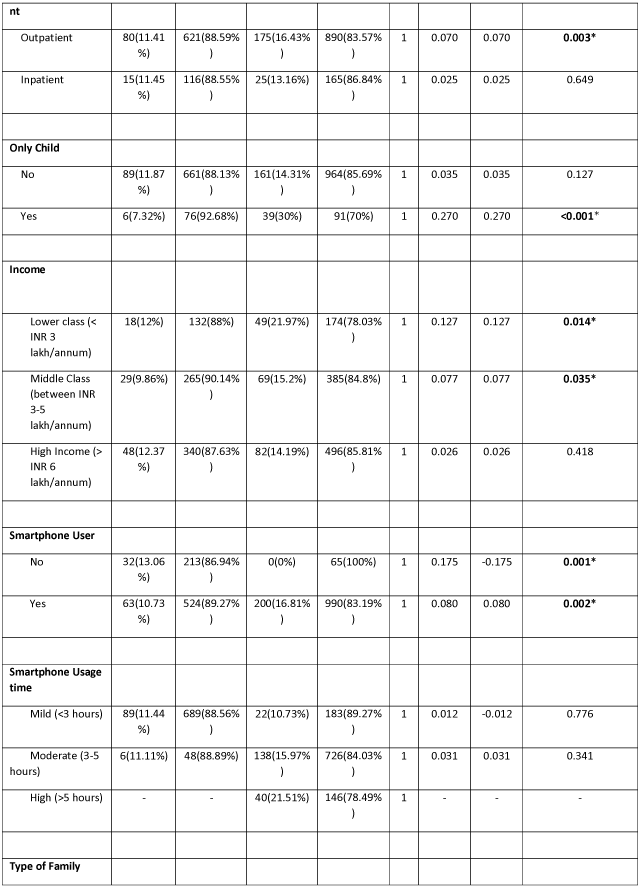
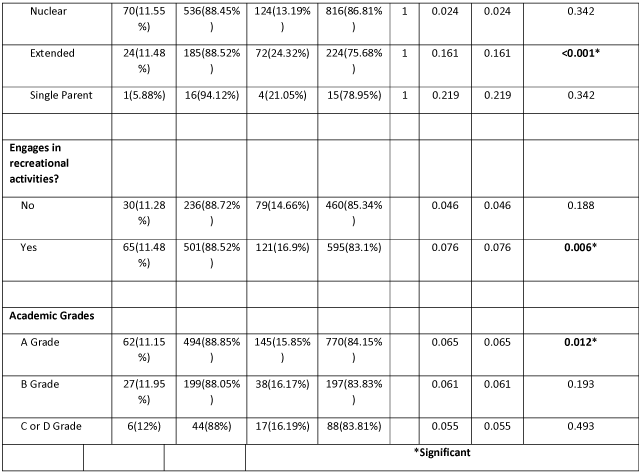
Depression using CES-DC before and after the COVID-19 lockdown by group.

### Factors associated with increased anxiety and depression during the pandemic

Regarding SCAS Scores, Mann-Whitney U and Kruskal–Wallis H test analyses showed that seven variables had a significant difference in anxiety levels (p<0.05), results of which are shown in Supplementary Table 2. The factors which showed increased anxiety levels during the pandemic included female gender (p<0.001), only child status (p=0.035), recreational activities (p<0.001), smartphone use (<0.001), friends or family members infected with COVID-19 (p<0.001), observing COVID-19 appropriate behaviour (p<0.001), and significant depressive symptoms using CES-DC (p<0.001). Additionally, we conducted a multiple linear regression model to analyse further the significant factors associated with children’s anxiety levels. We obtained the following factors to construct a multiple linear regression model of anxiety levels: Female gender (p<0.001, β coefficient = 0.491), extended family (p<0.001, β coefficient = 0.111), single-parent family (p=0.001, β coefficient = 0.042), smartphone use duration (p=<0.001, β coefficient = 0.103), family member or friend infected with COVID-19 (p<0.001, β coefficient = 0.210), observing COVID-19 appropriate behaviour (p=<0.001, β coefficient = 0.247), and significant depressive symptoms using CES-DC (p<0.001, β coefficient = 0.416). Together, these variables explained 78.4% of the variance in the anxiety levels. (F = 652.035, p < 0.001, R^2^=0.785, Adjusted R^2^=0.784) (Table 4).

**Table 4:** Factors associated with the anxiety levels of respondents during the COVID-19 outbreak (N = 1255).

| Model | Unstandardised Coefficients |  | Standardised Coefficients Beta | t | p-value |
| --- | --- | --- | --- | --- | --- |
|  | B | Std. Error |  |  |  |
| (Constant) | 3.480 | 0.654 |  | 5.320 | <0.001* |
| Significant depressive symptoms using CES-DC | 0.673 | 0.025 | 0.416 | 26.469 | <0.001* |
| Female Gender | 13.619 | 0.385 | 0.491 | 35.366 | <0.001* |
| Observing COVID-19 appropriate behaviour | 8.536 | 0.464 | 0.247 | 18.396 | <0.001* |
| Friend or Family member infected with COVID-19 | 5.963 | 0.414 | 0.210 | 14.387 | <0.001* |
| Extended Family type | 3.620 | 0.436 | 0.111 | 8.299 | <0.001* |
| Smartphone Usage Hours | 0.917 | 0.118 | 0.103 | 7.770 | <0.001* |
| Single parent family | 4.771 | 1.490 | 0.042 | 3.202 | 0.001* |
| F = 652.035, p < 0.001, R<sup>2</sup>=0.785, Adjusted R<sup>2</sup>=0.784. |  |  |  |  |  |
| **\*Significant** |  |  |  |  |  |
| Note: Forward stepwise selection procedure was employed to select the multiple linear model. |  |  |  |  |  |

To evaluate the factors associated with having depressive symptoms in children, binary logistic regression analysis was performed. We identified six factors as being significantly associated with increased levels of children’s clinical depressive symptoms: only-child status (p<0.001, OR=10.456), extended family (p<0.001, OR=2.754), family members or friends infected with COVID-19 (p<0.001, OR=89.571), a family member or friend died due to COVID-19 (p=0.032, OR=5.016), observing COVID-19 appropriate behaviour (p<0.001, OR=73.763), higher SCAS scores (p<0.001, OR=1.053), and smartphone usage duration (p=0.029, OR=1.186). However, no factor was significantly associated with decreased levels of children’s clinical depressive symptoms. (Table 5).

**Table 5:**
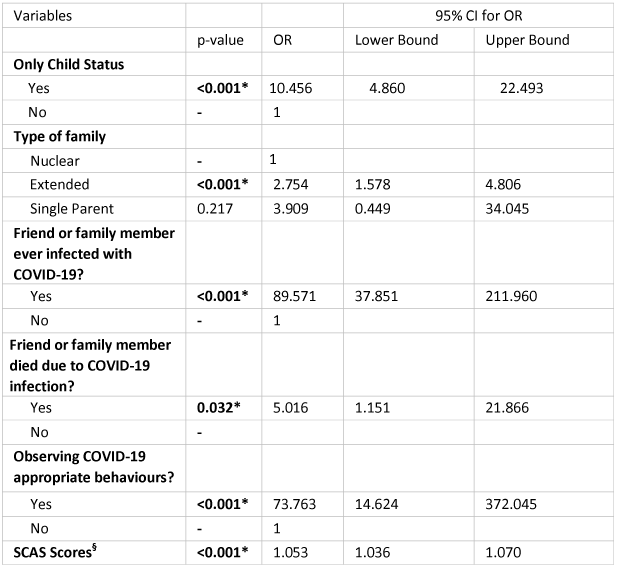
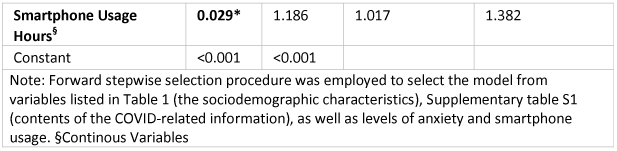
Factor associated with increased levels of clinical depressive symptoms of children in the COVID-19 pandemic. (N=1255)

| Variables |  |  | 95% CI for OR |  |
| --- | --- | --- | --- | --- |
|  | p-value | OR | Lower Bound | Upper Bound |
| **Only Child Status** |  |  |  |  |
| Yes | **<0.001\*** | 10.456 | 4.860 | 22.493 |
| No | - | 1 |  |  |
| **Type of family** |  |  |  |  |
| Nuclear | - | 1 |  |  |
| Extended | **<0.001\*** | 2.754 | 1.578 | 4.806 |
| Single Parent | 0.217 | 3.909 | 0.449 | 34.045 |
| **Friend or family member ever infected with COVID-19?** |  |  |  |  |
| Yes | **<0.001\*** | 89.571 | 37.851 | 211.960 |
| No | - | 1 |  |  |
| **Friend or family member died due to COVID-19 infection?** |  |  |  |  |
| Yes | **0.032\*** | 5.016 | 1.151 | 21.866 |
| No | - |  |  |  |
| **Observing COVID-19 appropriate behaviours?** |  |  |  |  |
| Yes | **<0.001\*** | 73.763 | 14.624 | 372.045 |
| No | - | 1 |  |  |
| **SCAS Scores<sup>§</sup>** | **<0.001\*** | 1.053 | 1.036 | 1.070 |
| **Smartphone Usage Hours<sup>§</sup>** | **0.029\*** | 1.186 | 1.017 | 1.382 |
| Constant | <0.001 | <0.001 |  |  |
| Note: Forward stepwise selection procedure was employed to select the model from variables listed in Table 1 (the sociodemographic characteristics), Supplementary table S1 (contents of the COVID-related information), as well as levels of anxiety and smartphone usage. §Continuous Variables |  |  |  |  |

### Child Abuse

113 (9%) parents reported beating their children during the pandemic, and 335 (26.7%) parents reported yelling at them.

## DISCUSSION

The present study on the long-term (>1 year) mental health impact of the COVID-19 pandemic and associated lockdown on children and adolescents is one of the few studies from low-to-middle income countries. There have been quite a few studies in developed countries regarding the long-term impact and fluctuation in children’s mental health symptoms during the pandemic,[35–38]. In this study, we found that levels of anxiety and depressive symptoms increased considerably during the pandemic. We also found that the prevalence of smartphone usage increased significantly during the pandemic. Only-child status, extended family, family members or friends infected with COVID-19, a family member or friend died due to COVID-19 infection, observing COVID-19 appropriate behaviour, higher SCAS scores, and smartphone usage duration were the factors associated with increased levels of depressive symptoms. No factor was significantly associated with decreased levels of children’s clinical depressive symptoms. Increased anxiety levels were associated with female gender, extended family, single-parent family, higher smartphone use duration, family member or friend infected with COVID-19, observing COVID-19 appropriate behaviour, and significant depressive symptoms using CES-DC.

The prevalence of depressive symptoms in our study rose from 11% pre-lockdown to 16% during the lockdown. In the English National survey follow-up, mental health problems, including depression, rose from 10.8% in 2017 to 16.0% in July 2020,[37]. In the continuation of this survey in the second wave, the prevalence of mental health disorders further increased to 17.4%,[36]. In another nationwide study in Germany, depression and other mental health issues increased from 9.9% before the pandemic to 17.8% during the pandemic,[39]. In other studies conducted during the lockdown, the prevalence of depressive symptoms in children and adolescents varied from 18% to 41% during the pandemic,[15,40,41]. Many cross-sectional studies conducted during the pandemic revealed an increased incidence of anxiety during the lockdown, consistent with our findings,[15,42– 44]. However, they compared the prevalence to studies conducted before the pandemic,[15,42,43].

Our findings suggest that the prevalence of smartphone use increased significantly during the pandemic. This could be attributed to the children’s social media use, video games, online schooling, lectures, and digital homework. Many of the studies conducted during the lockdown period have reported the association of smartphone use and internet addiction with poor mental health,[24,45–47], in line with our study. Smartphone use leads to unhealthy behaviours such as sedentary behaviour, a reduction in time dedicated to academic learning, and the replacement of all other forms of social relations with the smartphone (favouring a state of isolation and a tendency towards introversion),[48–51]. These factors were exacerbated during the pandemic, most likely contributing to increased mental health problems. On the other hand, smartphones offer the advantage of allowing youngsters to communicate with their acquaintances in real-time, promoting socialisation while also delivering the benefit of distance schooling. Children may acquire information via online resources such as encyclopaedias, internet searches, dictionaries, and educational applications. The benefits of smartphones cannot be overstated, and with these balanced benefits and drawbacks in mind, optimal usage of smartphones by children with parental supervision should be encouraged.

Surprisingly, 9% of the parents reported beating their children, and 25% reported yelling at them. It is thought that the pandemic has increased children’s exposure to violence in their homes and communities and hampered child safety services’ ability to recognise and respond to cases of abuse,[52]. Throughout the COVID-19 pandemic, however, police and social service agencies have noted a decrease in reporting of such claims. School closures, which kept children at home, may have aggravated these problems,[53]. The constant interaction between perpetrators and victims might have increased the violence and made reporting difficult. Preventative measures and assistance programs are required to address these challenges.

The majority of earlier studies were conducted exclusively as online surveys during the early stages of the pandemic because social interaction was restricted during the lockdown,[14,20,24,31,54]. These studies are less credible than traditional offline studies,[55] in which trained investigators interview participants one-on-one. These online studies are more prone to a substantial sampling bias due to non-probability sampling in which those who are more often in need are excluded,[56]. Furthermore, these online surveys have primarily included the adolescent age groups rather than children. On the contrary, in our study, children were more than adolescents [median age 10 (4) years]. The repeated cross-sectional study design allowed us to compare anxiety and depression levels in children and adolescents before and after the COVID-19 lockdown and school closures. Our study used data from the same settings before the pandemic, removing any confounding factors. Apart from these, we used two established and scientifically sound questionnaires in our study, making it one of the strengths. Every study has its own set of limitations, and ours is no exception. Because the data was collected in a hospital setting, not all of the individuals included in the study were healthy. The children who participated were visiting the hospital the mild sickness, immunisation or refilling of medications. Enrolling healthy children was impractical because schools and playgroups were closed, and we wanted to conduct an offline study, considering the sampling limitations of the other online studies. There was no way to overcome this limitation, so we included the available population. Another major limitation of the study is the lack of psychometric validation and the subjective nature of the COVID-19 pandemic-related questionnaire. Studies are necessary to develop and validate such questionnaires, which assess the general status of children within the context of the pandemic. Moreover, our study did not have a pre-considered hypothesis or study design, and there was an ad-hoc change in the pandemic according to the circumstances. However, because we compared this data to children who visited the same settings before the pandemic, we believe that these biases were eliminated. Also, CES-DC assesses the depressive symptoms in the past week, and as many of the children were attending hospital, the underlying disease may have caused the depressive symptoms in the past week.

Despite these limitations, this study provides invaluable information on the psychological status of the children in the worst-affected city in India more than a year after the outbreak of COVID-19. Our findings could serve as a historical reference. Most importantly, our findings highlight the need for psychological interventions to reduce psychological impact, anxiety, depression, and stress during the COVID-19 pandemic, as well as provide a baseline for evaluating prevention, control, and treatment efforts by all appropriate (government and non-government) agencies for the remainder of the pandemic.

We were unable to perform a longitudinal study that would offer clear evidence of the incidence of anxiety and depression during the study period, as well as the necessity for targeted public health approaches and interventions required for the well-being of the youngsters. A longitudinal study of this type may be developed to assess changes at the individual level. Additionally, the long-term effects of COVID-19 lockdown on other psychological aspects such as sleep disturbances, loneliness, and so on are unknown. Studies focusing on these aspects can be designed in the future.

## CONCLUSION

Many children experienced increased anxiety and depression due to the 15-month-long pandemic, lockdown, and school closures that followed, suggesting the need for interventions that engage children in healthy habits that protect children’s mental health and for ongoing monitoring of children in such situations to help prevent future mental health problems. Children’s increased smartphone use in the lockdown was more associated with psychosocial problems, so children and parents should be advised to encourage children to limit smartphone use. Only-child status, extended family, family members or friends infected with COVID-19, a family member or friend died due to COVID-19 infection, observing COVID-19 appropriate behaviour, higher SCAS scores, and smartphone usage duration were the factors associated with increased levels of depressive symptoms. No factor was significantly associated with decreased levels of children’s clinical depressive symptoms. Higher anxiety levels were associated with female gender, extended family, single-parent family, higher smartphone use duration, family member or friend infected with COVID-19, observing COVID-19 appropriate behaviour, and significant depressive symptoms using CES-DC. Our findings can help develop psychological interventions for the promotion and resilience of mental health in the ongoing pandemic and similar future conditions.

## Data Availability

All data produced in the present study are available upon reasonable request to the authors

---

**Supplementary Figure 1:** a) Violin plots of SCAS scores for Children (median (solid square)) b) Violin plots of SCAS scores for Adolescents (median (solid square)); c) Violin plots of SCAS scores for all participants (median (solid square)); d) Violin plots of CES-DC for Children (median (solid square)); e) Violin plots of CES-DC scores for adolescents (median (solid square)); f) Violin plots of CES-DC scores for all participants (median (solid square)); P-values show the SCAS/CES-DC scores significantly differ before and during the COVID-19 pandemic using Mann-Whitney U Test. *Significant

## Ethics Committee Clearance

Obtained. Date obtained: 31/03/2021

## IEC Clearance Number

IEC/Pharm/RP/387/Mar/2021

## Ethics Committee

Institutional Ethics Committee, Grant Govt. Medical College & Sir JJ Group of Hospitals, Byculla, Mumbai – 400008

## Acknowledgements

We would like to thank the Department of Public Health and Statistics of Grant Govt. Medical College and Ankush Jadhav from the Department of Biostatistics, NDMVP College Of Pharmacy, Nashik, for their expertise and assistance with the study’s statistical analysis. We would also like to thank the Department of Psychiatry and Department of Pharmacology, Grant Government Medical College and Sir JJ Group of Hospitals, Mumbai for their guidance and help. We also would like to thank the Indian Council of Medical Research, for guiding the students through their short-term studentship program.

## Data Availability Statement

The data that support the findings of this study are available from the corresponding author upon reasonable request.

## Contribution Details

Conceptualisation, MP and SM.; methodology, MP, SM, and MAG; Data collection, MP, SM, MAG, SAM, MS, VM, ZM, AM, AC, KD, SP, AJ, SW, DV, AB validation, MP, SM, MAG, SAM, MS, VM, ZM, AM, AC, KD, SP, AJ, SW, DV, AB.; formal analysis MP, SM, MAG, SAM, MS, VM, ZM, AM, AC, KD, AB.; data curation.; writing—original draft preparation, MP, SM, and MAG; writing—review and editing, MP, SM, MAG, SAM, MS, VM, ZM, AM, AC, KD, SP, AJ, SW, DV, AB; supervision, MP, SM, and MAG.; project administration, MP, SM and MAG. All authors have read and agreed to the published version of the manuscript.

## REFERENCES

1 Kim C, Ko H. The impact of self-compassion on mental health, sleep, quality of life and life satisfaction among older adults. Geriatr Nurs 2018;39:623–8. doi:10.1016/j.gerinurse.2018.06.005

2 Covid-19: India school closures “catastrophic” for poor students - BBC News. https://www.bbc.com/news/world-asia-india-58432648 (accessed 11 Oct 2021).

3 Education: From disruption to recovery. UNESCO. 2020.https://en.unesco.org/covid19/educationresponse (accessed 15 Feb 2022).

4 Chen S. Psychological adjustment during the global outbreak of COVID-19: A resilience perspective. Psychological Trauma: Theory, Research, Practice, and Policy 2020 0615;12:S51. doi:10.1037/tra0000685

5 Covid-19: India school closures “catastrophic” for poor students. BBC News. 2021.https://www.bbc.com/news/world-asia-india-58432648 (accessed 15 Feb 2022).

6 Brooks SK, Webster RK, Smith LE, et al. The psychological impact of quarantine and how to reduce it: rapid review of the evidence. Lancet 2020;395:912–20. doi:10.1016/S0140-6736(20)30460-8

7 Maharashtra COVID lockdown: Offices, local trains, cinema halls — what’s allowed, what’s not. https://www.livemint.com/news/india/maharashtra-covid-lockdown-offices-transport-hotel-what-s-allowed-what-s-not-11617547148962.html (accessed 2 Feb 2022).

8 Pfefferbaum B. Challenges for Child Mental Health Raised by School Closure and Home Confinement During the COVID-19 Pandemic. Curr Psychiatry Rep 2021;23:65. doi:10.1007/s11920-021-01279-z

9 Yeasmin S, Banik R, Hossain S, et al. Impact of COVID-19 pandemic on the mental health of children in Bangladesh: A cross-sectional study. Child Youth Serv Rev 2020;117:105277. doi:10.1016/j.childyouth.2020.105277

10 Fegert JM, Vitiello B, Plener PL, et al. Challenges and burden of the Coronavirus 2019 (COVID-19) pandemic for child and adolescent mental health: a narrative review to highlight clinical and research needs in the acute phase and the long return to normality. Child Adolesc Psychiatry Ment Health 2020;14:20. doi:10.1186/s13034-020-00329-3

11 Kendler KS, Karkowski LM, Prescott CA. Causal relationship between stressful life events and the onset of major depression. Am J Psychiatry 1999;156:837–41. doi:10.1176/ajp.156.6.837

12 Adverse consequences of school closures. UNESCO. 2020.https://en.unesco.org/covid19/educationresponse/consequences (accessed 15 Feb 2022).

13 Scobie G, Whitehead R. What are the impacts of past infectious disease outbreaks on non-communicable health outcomes. Edinburgh: Public Health Scotland 2020;:3.

14 Zhou S-J, Zhang L-G, Wang L-L, et al. Prevalence and sociodemographic correlates of psychological health problems in Chinese adolescents during the outbreak of COVID-19. Eur Child Adolesc Psychiatry 2020;29:749–58. doi:10.1007/s00787-020-01541-4

15 Tang S, Xiang M, Cheung T, et al. Mental health and its correlates among children and adolescents during COVID-19 school closure: The importance of parent-child discussion. J Affect Disord 2021;279:353–60. doi:10.1016/j.jad.2020.10.016

16 Racine N, McArthur BA, Cooke JE, et al. Global Prevalence of Depressive and Anxiety Symptoms in Children and Adolescents During COVID-19: A Meta-analysis. JAMA Pediatr 2021;175:1142–50. doi:10.1001/jamapediatrics.2021.2482

17 Robinson E, Sutin AR, Daly M, et al. A systematic review and meta-analysis of longitudinal cohort studies comparing mental health before versus during the COVID-19 pandemic in 2020. J Affect Disord 2022;296:567–76. doi:10.1016/j.jad.2021.09.098

18 Orgilés M, Morales A, Delvecchio E, et al. Immediate Psychological Effects of the COVID-19 Quarantine in Youth From Italy and Spain. Front Psychol 2020;11:579038. doi:10.3389/fpsyg.2020.579038

19 Garcia de Avila MA, Hamamoto Filho PT, Jacob FL da S, et al. Children’s Anxiety and Factors Related to the COVID-19 Pandemic: An Exploratory Study Using the Children’s Anxiety Questionnaire and the Numerical Rating Scale. Int J Environ Res Public Health 2020;17:E5757. doi:10.3390/ijerph17165757

20 Jiao WY, Wang LN, Liu J, et al. Behavioral and Emotional Disorders in Children during the COVID-19 Epidemic. J Pediatr 2020;221:264–266.e1. doi:10.1016/j.jpeds.2020.03.013

21 Singh S, Roy D, Sinha K, et al. Impact of COVID-19 and lockdown on mental health of children and adolescents: A narrative review with recommendations. Psychiatry Res 2020;293:113429. doi:10.1016/j.psychres.2020.113429

22 Saurabh K, Ranjan S. Compliance and Psychological Impact of Quarantine in Children and Adolescents due to Covid-19 Pandemic. Indian J Pediatr 2020;:1–5. doi:10.1007/s12098-020-03347-3

23 Pinar Senkalfa B, Sismanlar Eyuboglu T, Aslan AT, et al. Effect of the COVID-19 pandemic on anxiety among children with cystic fibrosis and their mothers. Pediatr Pulmonol 2020;55:2128–34. doi:10.1002/ppul.24900

24 Duan L, Shao X, Wang Y, et al. An investigation of mental health status of children and adolescents in china during the outbreak of COVID-19. J Affect Disord 2020;275:112–8. doi:10.1016/j.jad.2020.06.029

25 Colizzi M, Sironi E, Antonini F, et al. Psychosocial and Behavioral Impact of COVID-19 in Autism Spectrum Disorder: An Online Parent Survey. Brain Sci 2020;10:E341. doi:10.3390/brainsci10060341

26 The DEPRESSD Project. DEPRESSD Project. https://www.depressd.ca/covid-19-mental-health (accessed 7 Feb 2022).

27 Spence SH. A measure of anxiety symptoms among children. Behav Res Ther 1998;36:545–66. doi:10.1016/s0005-7967(98)00034-5

28 Omkarappa DB, Rentala S. Anxiety, depression, self-esteem among children of alcoholic and nonalcoholic parents. J Family Med Prim Care 2019;8:604–9. doi:10.4103/jfmpc.jfmpc_282_18

29 Spence SH, Barrett PM, Turner CM. Psychometric properties of the Spence Children’s Anxiety Scale with young adolescents. J Anxiety Disord 2003;17:605–25. doi:10.1016/s0887-6185(02)00236-0

30 Karande S, Gogtay N, Bala N, et al. Anxiety symptoms in regular school students in Mumbai City, India. J Postgrad Med 2018;64:92–7. doi:10.4103/jpgm.JPGM_445_17

31 Orgilés M, Fernández-Martínez I, Guillén-Riquelme A, et al. A systematic review of the factor structure and reliability of the Spence Children’s Anxiety Scale. Journal of Affective Disorders 2016;190:333–40. doi:10.1016/j.jad.2015.09.055

32 Faulstich ME, Carey MP, Ruggiero L, et al. Assessment of depression in childhood and adolescence: an evaluation of the Center for Epidemiological Studies Depression Scale for Children (CES-DC). Am J Psychiatry 1986;143:1024–7. doi:10.1176/ajp.143.8.1024

33 Betancourt T, Scorza P, Meyers-Ohki S, et al. Validating the Center for Epidemiological Studies Depression Scale for Children in Rwanda. J Am Acad Child Adolesc Psychiatry 2012;51:1284–92. doi:10.1016/j.jaac.2012.09.003

34 Center for Epidemiological Studies Depression Scale for Children (CES-DC). NovoPsych. 2021.https://novopsych.com.au/assessments/diagnosis/center-for-epidemiological-studies-depression-scale-for-children-ces-dc/ (accessed 18 Jan 2022).

35 Co-Space Study. https://cospaceoxford.org/ (accessed 15 Feb 2022).

36 Mental Health of Children and Young People in England 2021 - wave 2 follow up to the 2017 survey. NHS Digital. https://digital.nhs.uk/data-and-information/publications/statistical/mental-health-of-children-and-young-people-in-england/2021-follow-up-to-the-2017-survey (accessed 15 Feb 2022).

37 Newlove-Delgado T, McManus S, Sadler K, et al. Child mental health in England before and during the COVID-19 lockdown. Lancet Psychiatry 2021;8:353–4. doi:10.1016/S2215-0366(20)30570-8

38 Young People’s Mental Health during the COVID-19 Pandemic. NIHR School for Public Health Research. 2020.https://sphr.nihr.ac.uk/wp-content/uploads/2020/08/Young-Peoples-Mental-Health-during-the-COVID-19-Pandemic-Report-Final.pdf (accessed 18 Mar 2022).

39 Ravens-Sieberer U, Kaman A, Erhart M, et al. Impact of the COVID-19 pandemic on quality of life and mental health in children and adolescents in Germany. Eur Child Adolesc Psychiatry Published Online First: 25 January 2021. doi:10.1007/s00787-021-01726-5

40 Panda PK, Gupta J, Chowdhury SR, et al. Psychological and Behavioral Impact of Lockdown and Quarantine Measures for COVID-19 Pandemic on Children, Adolescents and Caregivers: A Systematic Review and Meta-Analysis. J Trop Pediatr 2021;67:fmaa122. doi:10.1093/tropej/fmaa122

41 Sama BK, Kaur P, Thind PS, et al. Implications of COVID-19-induced nationwide lockdown on children’s behaviour in Punjab, India. Child Care Health Dev 2021;47:128–35. doi:10.1111/cch.12816

42 Zhang L, Zhang D, Fang J, et al. Assessment of Mental Health of Chinese Primary School Students Before and After School Closing and Opening During the COVID-19 Pandemic. JAMA Netw Open 2020;3:e2021482. doi:10.1001/jamanetworkopen.2020.21482

43 Xie X, Xue Q, Zhou Y, et al. Mental Health Status Among Children in Home Confinement During the Coronavirus Disease 2019 Outbreak in Hubei Province, China. JAMA Pediatr 2020;174:898–900. doi:10.1001/jamapediatrics.2020.1619

44 Qin Z, Shi L, Xue Y, et al. Prevalence and Risk Factors Associated With Self-reported Psychological Distress Among Children and Adolescents During the COVID-19 Pandemic in China. JAMA Netw Open 2021;4:e2035487. doi:10.1001/jamanetworkopen.2020.35487

45 Serra G, Lo Scalzo L, Giuffrè M, et al. Smartphone use and addiction during the coronavirus disease 2019 (COVID-19) pandemic: cohort study on 184 Italian children and adolescents. Ital J Pediatr 2021;47:150. doi:10.1186/s13052-021-01102-8

46 Shah S, Kaul A, Shah R, et al. Impact of Coronavirus Disease 2019 Pandemic and Lockdown on Mental Health Symptoms in Children. Indian Pediatr 2021;58:75–6.

47 Ren H, He X, Bian X, et al. The Protective Roles of Exercise and Maintenance of Daily Living Routines for Chinese Adolescents During the COVID-19 Quarantine Period. J Adolesc Health 2021;68:35–42. doi:10.1016/j.jadohealth.2020.09.026

48 González-Bueso V, Santamaría JJ, Fernández D, et al. Association between Internet Gaming Disorder or Pathological Video-Game Use and Comorbid Psychopathology: A Comprehensive Review. Int J Environ Res Public Health 2018;15:E668. doi:10.3390/ijerph15040668

49 Schuurmans AAT, Nijhof KS, Vermaes IPR, et al. A Pilot Study Evaluating “Dojo,” a Videogame Intervention for Youths with Externalizing and Anxiety Problems. Games Health J 2015;4:401–8. doi:10.1089/g4h.2014.0138

50 Fish MT, Russoniello CV, O’Brien K. The Efficacy of Prescribed Casual Videogame Play in Reducing Symptoms of Anxiety: A Randomized Controlled Study. Games Health J 2014;3:291–5. doi:10.1089/g4h.2013.0092

51 Schuurmans AAT, Nijhof KS, Engels RCME, et al. Using a Videogame Intervention to Reduce Anxiety and Externalizing Problems among Youths in Residential Care: an Initial Randomized Controlled Trial. J Psychopathol Behav Assess 2018;40:344–54. doi:10.1007/s10862-017-9638-2

52 Bhatia A, Fabbri C, Cerna-Turoff I, et al. Violence against children during the COVID-19 pandemic. Bull World Health Organ 2021;99:730–8. doi:10.2471/BLT.20.283051

53 Kourti A, Stavridou A, Panagouli E, et al. Domestic Violence During the COVID-19 Pandemic: A Systematic Review. Trauma Violence Abuse 2021;:15248380211038690. doi:10.1177/15248380211038690

54 S T, M X, T C, et al. Mental health and its correlates among children and adolescents during COVID-19 school closure: The importance of parent-child discussion. Journal of affective disorders 2021;279. doi:10.1016/j.jad.2020.10.016

55 Ball HL. Conducting Online Surveys. J Hum Lact 2019;35:413–7. doi:10.1177/089033441984873456

56 Pierce M, McManus S, Jessop C, et al. Says who? The significance of sampling in mental health surveys during COVID-19. The Lancet Psychiatry 2020;7:567–8. doi:10.1016/S2215-0366(20)30237-6

